# Predicting Impulsive Choices: Development of a Novel Experimental Task

**DOI:** 10.64898/2026.03.11.26348147

**Authors:** Hongzhe Ma, Diede Fennema, Sara Simblett, Roland Zahn

## Abstract

**Aims:** Due to the multifaceted nature of “impulsivity”, its measurement remains fragmented. Here, we developed the Risky Social Choices task to provide evidence for its validity and reliability, while testing the hypothesis that impaired access to implicit knowledge of negative long-term consequences is of distinct importance for “impulsive” decision-making in a general population sample.

**Methods:** Forty participants chose whether to engage in risk-taking behaviors, which combined web-based AI-generated videos with narrated hypothetical scenarios and measured worries related to negative long-term consequences, approach-related motivation for short-term rewards, response time to and accuracy of recognizing degraded auditory prime words denoting negative long-term consequences.

**Results:** A pre-registered multi-step regression model was constructed with worry, motivation, response time and accuracy as predictors and percentage of risky choices as the outcome. Among all predictors, only prime word recognition accuracy was significantly negatively associated with risky choices, confirming our hypothesis of the role of reduced implicit access to negative long-term consequences in risk-taking decisions. In contrast, approach-related motivation for rewards was the only predictor significantly positively related to percentage of risky choices.

**Discussion:** As predicted, the negative association between risky choices and implicit access to negative long-term consequences supports its role as a distinct aspect of “impulsivity”. The novel task successfully captured this aspect, paving the way for a more precise neurocognitive characterization of clinical conditions where “impulsivity” plays a key role. The findings unveil the importance of implicit social sequential knowledge for impulsivity in neurotypical populations, so far only investigated in patients with brain lesions.

## Introduction

So-called “impulsivity” is a multifaceted construct central to a wide range of behaviors and clinical conditions, including substance/alcohol use disorders (SUD/AUD), attention-deficit/hyperactivity disorder (ADHD), mania/hypomania, and neurodegenerative diseases (American Psychiatric Association, 2013; Bakhshani, 2014; Dalley et al., 2011). Due to its complex nature, the definition of impulsivity has been discussed in multiple contexts, but its measurement remains fragmented. Self-report scales and behavioral tasks often show weak convergence, and different paradigms appear to capture distinct underlying mechanisms rather than a single, unified construct (Cyders & Coskunpinar, 2011). This lack of convergence thus presents a methodological challenge for impulsivity research, which is that researchers may label a certain behavior as impulsive while relying on measures that differ substantially in what they assess. Thus, specifying which aspect of impulsivity is being studied and matching that aspect of interest to the measure that actually captures the underlying cognitive process is essential for drawing more accurate conclusions about factors underpinning impulsive, risky, or socially inappropriate behaviors.

When viewed as a personality trait, impulsivity is described as relatively stable patterns of thoughts, feelings, and behaviors that reflect an individual’s tendency to act impulsively and distinguish them from others in the general population (Deyoung, 2010; Roberts & Mroczek, 2008). This focus on response patterns rather than a single act is commonly assessed as “trait impulsivity” in the clinical domain using the Barratt Impulsiveness Scale (BIS-11; (Patton et al., 1995)). Previous research has demonstrated trait impulsivity to be a significant predictor of several maladaptive behaviors, including substance use among university students, suicide attempts in patients with bipolar disorder, and binge eating behaviours in adolescents (Hamdan-Mansour et al., 2018; Lee-Winn et al., 2016; Swann et al., 2005). This is also consistent with findings that patients with high impulsivity traits, such as ADHD, are at higher risk of developing a substance use disorder or behavioral addiction (Crunelle et al., 2013), and that disorders characterized by impulsive response patterns often coexist (Grant & Kim, 2003).

In contrast to reflecting a stable predisposition, impulsivity can also be understood as a rash action that arises in the moment in response to immediate cues (Cyders & Coskunpinar, 2011). More specifically, impulsivity can manifest aspects like loss of motor or response “inhibition”, deficits in delaying rewards, impaired resistance to distractor interference, impaired resistance to proactive interference, and distortions in elapsed time. Behavioral tasks, such as the Go/No-Go task, Stroop test, and Eriken Flanker task, have been developed to test a participant’s ability to inhibit motor or cognitive responses as a measure of impulsivity (Bezdjian et al., 2009; Dimoska-Di Marco et al., 2011; Eriksen & Eriksen, 1974).

One dominant model conceptualizes impulsivity as a failure of inhibitory control, or an inability to inhibit behavioral impulses and thoughts (Bari & Robbins, 2013; Daruna & Barnes, 1993; Miller & Cohen, 2001). In line with this perspective, impulsivity is generally accepted as the tendency to act prematurely without foresight in psychopathology (Dalley et al., 2011).This cognitive control model of impulsivity essentially implies that responses occur before one can reflectively weigh the outcomes or consequences of one’s behavior, making the responses appear inappropriate or risky. This feature thus distinguishes impaired judgment from impulsivity, where one’s knowledge of the longer-term consequences of social behavior should be intact, although access or the motivational impact of it may be weakened. Consequently, an individual’s behavior will not be considered impulsive in the core sense when the individual’s long-term knowledge of the consequences of social behavior is impaired. A contrasting account, however, suggests that some behaviors labeled as “impulsive” may indeed arise from impaired access to knowledge about long-term negative consequences at the moment of decision-making. A previous study in patients with frontotemporal lobar neurodegeneration (FTLD) demonstrated that the patients’ seemingly impulsive or inappropriate social behavior was likely the result of impaired social knowledge of long-term negative consequences (Zahn et al., 2017), consistent with representational models proposing that the medial prefrontal cortex encodes structured representations of social event knowledge that guide behavior in complex social contexts (Krueger & Grafman, 2009). Although it is unlikely that the same knowledge is impaired in patients without brain lesions, it is possible they experience more difficulties accessing knowledge of the consequences of social behavior or evaluate negative consequences differently, rather than an inability to control inappropriate urges. Specifically, the motivational impact of anticipated long-term negative consequences may be relatively weak, while sensitivity to short-term rewards remains strong. When long-term consequences exert less influence on decision-making, behavior becomes disproportionately guided by the immediate motivational pull of short-term rewards. This imbalance would increase the likelihood of engaging in reward-driven actions despite potential future costs, or in other words, impulsive behavior may arise from an imbalance between competing motivational systems. That is, this mechanism would account for core aspects of impulsivity without resorting to an urge suppression model and instead consider competing motivations in a self-controlling frontal-temporal-subcortical network as proposed by previous work in the event-feature-emotion complex model (Moll et al., 2005; Zahn et al., 2020).

This distinction thus highlights an important gap in impulsivity research. Most existing tasks focus either on reward sensitivity or inhibitory control, but few directly examine the accessibility of consequences during decision-making. In addition, consequence accessibility and motivational factors are rarely measured within the same task. As a result, it is difficult to determine whether increased motivation for reward, diminished concern for outcomes, or reduced access to consequence knowledge underpins risky or socially inappropriate behavior. A method that integrates these components within a single task could therefore offer a clearer picture of the cognitive processes behind impulsive decision-making.

The current study introduces a novel experimental task named the Risky Social Choices (RSC) to address this gap. In this task, participants are presented with binary choices in hypothetical social scenarios where they have to decide whether to carry out a socially inappropriate or risky act. Each decision is immediately followed by a word recognition task, where the participants attempt to recognize an aurally degraded word stating the long-term negative consequence of that impulsive act. Participants then view a short outcome video depicting the consequences of the chosen action and provide ratings of anticipated motivational state and worry towards future outcome. Their reaction time and accuracy in recognizing the degraded words are used as implicit measures of accessibility of long-term negative consequences. Implicit measurement strategies can potentially offer insights into a cognitive domain that are not reachable through self-report measures (Greenwald et al., 2002). In this case, the role of long-term negative consequences of social behaviour is assessed implicitly via a phenomenon called priming. Priming is an unconscious memory process that enhances a person’s ability to recognize generate, or categorize an item due to prior exposure to that item or something similar (Schacter et al., 2004). When used as an implicit measure to probe whether an individual processes certain knowledge, the priming effect can be evident from the individual’s faster response time or higher accuracy in recognizing a target stimulus (Scarborough et al., 1977; Schacter & Buckner, 1998). Thus, theoretically speaking, when an individual has implicit access to the knowledge of long-term negative consequences of social behavior, it will be easier for that individual to recognize a long-term negative consequence, or something related to that negative consequence. Overall, by assessing implicit access to knowledge of long-term negative consequences, approach-related motivation for reward, and worry towards future negative consequences of impulsive behavior within a single task, the RSC task allows the investigation of a potential new aspect of impulsivity, which is weakened accessibility of knowledge of long-term negative consequences, and the role of multiple cognitive factors in impulsive decision-making.

The primary aim of this study was to evaluate the validity, reliability, and feasibility of this novel experimental task in a non-clinical online sample. Based on prior theoretical work and relevant studies, we hypothesized that a higher percentage of impulsive choices would be associated with lower degrees of worry related to negative long-term consequences, higher approach-related motivation for positive short-term consequences, longer response times to recognize negative consequence-related degraded auditory prime words, and lower accuracy in recognizing negative consequence-related degraded auditory prime words.

## Methods

### Participants

Participants were recruited online via Amazon Mechanical Turk and were compensated 8 pounds for their time upon completion of the online experimental task on Qualtrics. Participants provided written informed consent online in accordance with a King’s College London PNM Ethics Committee approved protocol (reference number: HR-23/24-41651) before participating in the study. No identifiable information was collected from the participants, and participants were allowed to drop out of the study at any time point without giving a reason.

All participants were UK residents, at least 18 years old, with sufficient English proficiency, normal or corrected-to-normal vision, no hearing loss or impairment, and had access to a computer or tablet to watch videos and hear narration. Participants who were unable to consent to the study, who had a drug or alcohol use condition that could be easily triggered by cues, specifically video clips depicting drug or alcohol use scenes, or who had severe distress or self-harm tendencies that could be easily triggered by sexual or violent depictions were excluded from the study (self-selected exclusion).

Sixty-three people participated in the study. After excluding participants with incomplete submissions, submissions with repeated responses to open-ended questions, and submissions with responses unrelated to the question prompts, a total of 40 participants were included in the final analysis.

### Task Overview

The study was a pre-registered cross-sectional study in a general population sample (Open Science Framework link: https://osf.io/fe5gp) designed to investigate the cognitive components underpinning impulsive decision-making using a novel experimental task called the RSC task (available under a Creative Commons – Attribution – CC BY licence without need to ask for permission). In the pre-registration, study design, task template, primary hypotheses, planned sample size, inclusion and exclusion criteria, primary variables, and analytic approach were all specified. All data generated or analyzed during this study can also be found in the above OSF article.

After providing their basic demographic information, participants were asked to complete BIS-11ro measure trait impulsivity and potentially validate the RCS task against it. Before the actual task began, each participant would go through two sets of practice questions to familiarize themselves with what to expect during the task. The actual task consisted of 44 sets of questions, and the entire study took approximately 40 minutes to complete.

Each set of questions started with one sentence of a narrated hypothetical risky scenario followed by a question asking the participant whether they would like to carry out an impulsive act. The participant was asked to imagine experiencing the situation in a first-person point-of-view and answer the question by making a binary “yes or no” choice, indicating whether they would like to carry out a specific impulsive act. After choosing an answer, the participant went on to the next question, where a clip of a degraded audio stimulus of a long-term consequence of the impulsive choice was played automatically. On the same page, the participant was asked to type down what they heard from the degraded audio stimulus. Based on what the participant chose for the first question, a 4-second AI-generated video was played. If the participant chose “yes” for the first question, a potential outcome of that act would be played; while if the participant chose “no” for the first question, a neutral control of an everyday object or scene would be played. The last question comprised two 5-point scales asking the participant to rate their reaction towards the risky scenario, specifically how much they would want to do something impulsive and how worried they would be about the long-term negative consequence of that impulsive act. A schematic example of a full set of questions is demonstrated in Figure 1 below.

**Figure 1.**
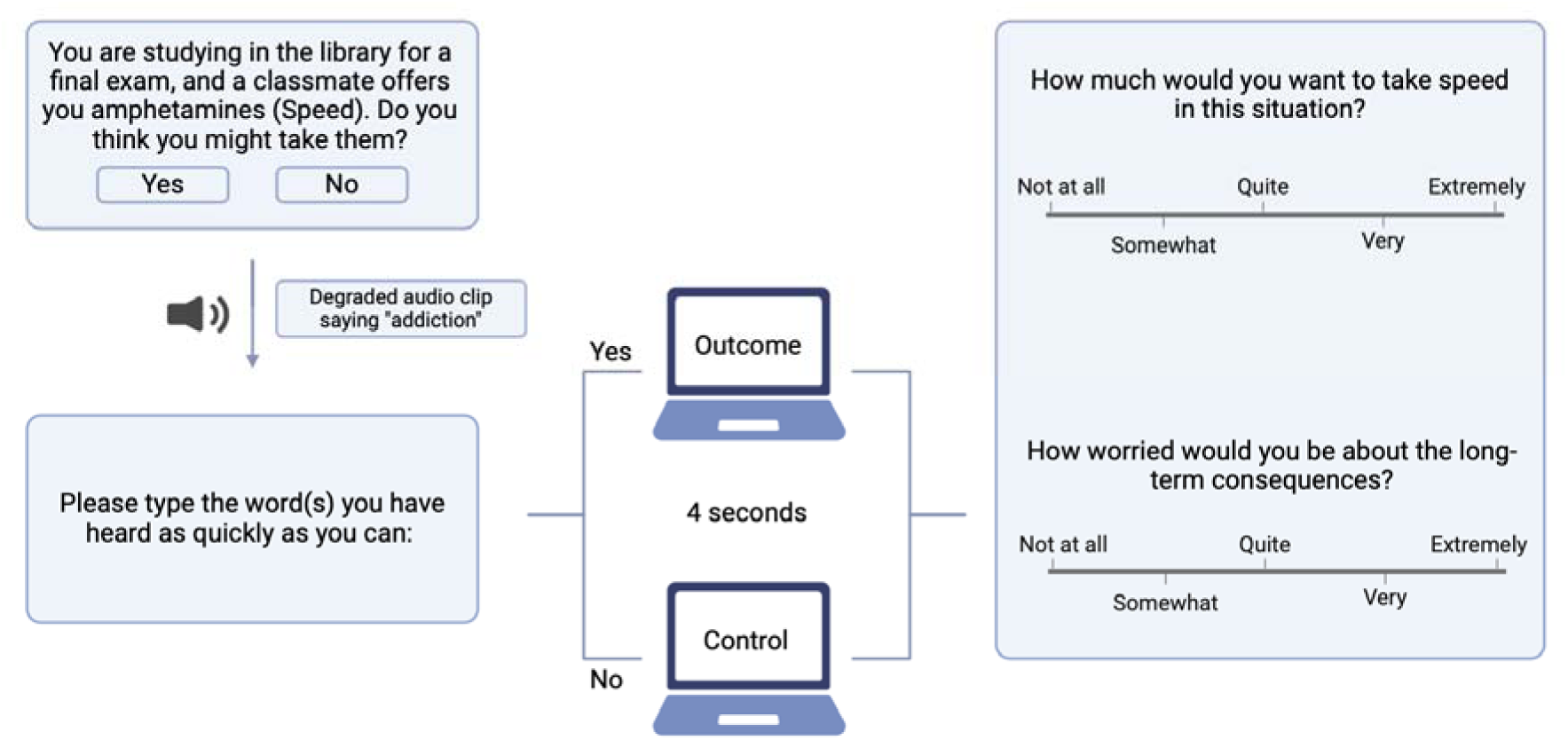
A demonstration of one set of questions in an example hypothetical risky scenario. Figure created using BioRender.com.

### Task Design

#### Risky Scenarios

To better understand impulsivity and investigate what factors underpin impulsive choices in an experimental setting, it is necessary to identify situations where people tend to make impulsive choices or situations where behavior may potentially lead to both a short-term reward and a negative long-term consequence. One defining characteristic of risky activities is their potential to cause negative outcomes or harm to the individual as well as the possibility of resulting in a reward or positive outcome (Leigh, 1999; MacPherson et al., 2010). Therefore, by creating hypothetical risky scenarios, researchers may be able to mimic real-life situations where people face the choice of whether or not to carry out certain impulsive acts.

A total of 44 hypothetical risky social scenarios where people tend to make impulsive decisions were created based on 30 risky activities identified in a previous study (Fromme et al., 1997) and 58 impulsive social situations collected by a past normative study (Zahn et al., 2017). These 44 scenarios generally fit into 10 categories, namely illicit drug use, aggressive behaviors, risky sexual activities, heavy drinking, high risk sports, academic/ work behaviors, overspending, binge eating, dating, and gambling. For example, one scenario could be “You are not satisfied with your current job. Do you think you might quit without having plans for your next job?”. Another example could be “You are at a casino and think you could win back the money you lost. Do you think you might play another round?”. Scenarios were designed to be understandable without specialized knowledge. The same set of scenarios was presented to all participants with randomized order across individuals.

#### Choice Response

Participants indicated their choice by selecting either “Yes” or “No” after hearing the narration of each scenario. Endorsement of the risky or socially inappropriate action (Yes) was accounted for as one risky choice made. No time limit was imposed on responses, but response times were recorded automatically on Qualtrics.

#### Negative Long-term Consequences

Eleven categories of negative long-term consequences were derived from the 44 hypothetical risky scenarios identified earlier. The consequences were addiction, criminal record, damaged reputation, traumatized, unplanned parenthood, overweight, chronic illness, permanent injury, unemployment, bankruptcy, and unhappiness.

#### Degraded Audio Stimuli

The 11 negative consequences, as well as the 44 risky scenarios, were transformed from text to sound using the AI voice generation software *ElevenLabs* (ElevenLabs, 2023). An AI voice called Lily, which was the voice of a young female with a British accent, was selected. Although worded rather abstractly, some of the risky scenarios referred to illegal, sexual, or violent activities. Thus, by having a narration of these scenarios and asking questions instead of using only words on a screen, more complete reporting of sensitive behavior was allowed (O’Reilly et al., 1994). All 11 negative consequences were then distorted using Audacity (Version 3.5.0), an audio editing software, by going through distortion effect -100 clipping three times to ensure the same amount of distortion. In this way, the explicit comprehensibility of these words was reduced to make sure prime words recognition relied more on spontaneous implicit judgement. Next, the sound volume of a one-second “whoosh” sound obtained from *Pixabay* (Pixabay, 2024) was lowered using Audacity by applying a −16 dB amplification. Lastly, the “whoosh” sound was added 0.2 second before the onset of each degraded consequence to direct participants’ attention to the audio stimulus.

#### AI-generated Video Clips

The text-to-video AI *Gen-2* (Runway, 2023) was used to generate all outcome video clips for the task. Each clip lasted 4 seconds long, depicting the potential outcome of the specific impulsive act from a first-person point-of-view to add more realism. However, these clips would only be displayed if the participant had chosen to carry out the impulsive act. Since some of the risky scenarios involved sexual indiscretion, what was displayed in the video clip depended on what each participant had chosen as their sexuality previously. If a participant chose not to engage in the risky activity, a neutral video clip also generated by the same AI would be displayed. The neutral videos were generally everyday objects, buildings, ordinary natural sceneries, and everyday facilities since they were shown to elicit neutral emotions in people (Li et al., 2022).

The video was played after the prime word question because it might potentially remind the participant of the long-term negative consequence of their decision and thus influence the accuracy of their prime word recognition. At the same time, it was placed before reactivity ratings (motivation and worry of carrying out an impulsive act) as a reinforcement or reminder of the risky scenario and the choice made.

### Measures

#### Demographic Information

Participants’ basic demographic information about their age, sex assigned at birth, gender, ethnic group/background, years of education, and sexuality was collected before the task began. In particular, sexuality was asked by presenting the participants with two artificial-intelligence (AI) -generated characters, one with more traditionally feminine features and the other one with more traditionally masculine features, and asking the participants which character they were more attracted to. This approach was adopted to capture sexuality as a behavioral preference rather than an identity label, reducing potential ambiguity in self-labeling or discomfort with explicit identity-based questions.

#### Barratt Impulsiveness Scale (BIS-11)

BIS-11 is one of the most frequently used self-report instruments in impulsivity research and is widely used to measure trait impulsivity (Patton et al., 1995; Reise et al., 2013). Past research has demonstrated BIS-11to be valid across different cultural and clinical samples (Patton et al., 1995; Vasconcelos et al., 2012) with acceptable to good internal consistency as indicated by Cronbach’s alphas ranging from 0.79 to 0.83 (Patton et al., 1995). The questionnaire consists of 30 items aiming to measure impulsiveness based on 6 first-order factors, including attention, motor, self-control, cognitive complexity, perseverance, and cognitive instability impulsiveness, which in turn form 3 second-order factors, including attentional, motor, and non-planning impulsiveness. Each item is rated on a 4-point scale, with 1 being “rarely/never” and 4 being “almost always/always”. The total score ranges from 30 to 120 with higher scores indicating higher levels of impulsiveness, and a score of 72 or above is generally considered highly impulsive.

#### Reactivity to Risky Scenarios

At the end of each scenario, participants were asked to rate approach-related motivation for positive short-term consequences by indicating on a five-point scale how much they would want to carry out specific impulsive acts in given situations, with 0 being “not at all” and 4 being “extremely”. They were also asked to rate the degree of worry related to negative long-term consequences by indicating on a five-point scale how worried they would be about the long-term consequences, with 0 being “not at all” and 4 being “extremely”.

#### Response Time and Accuracy

During the task, participants were asked to type down as soon as possible what they heard from a clip of aurally degraded word(s), which served as the prime word, in each risky scenario. Prime word recognition accuracy was categorized into no response, partially accurate, and fully accurate. The prime words were chosen to reflect long-term negative consequences to probe implicit access to knowledge of long-term consequences (Zahn et al., 2017). Participants’ response time to prime word in each scenario was also recorded, which was the overall time (in seconds) they spent understanding the degraded audio stimulus and typing down the response on the prime word recognition page.

### Statistical Analysis

All data was organized and analyzed using Microsoft Excel and IBM SPSS Statistics (Version 28.0.1.1). Choice responses were coded as 1 (Yes) and 0 (No) and percentage of risky choices, which was the main outcome variable in subsequent analyses, was computed as the percentage of “Yes” responses across all 44 scenarios. Accuracy in recognizing degraded auditory word stimuli was re-coded from “no response”, “partially accurate”, and “fully accurate” to 0, 1, and 2 respectively. All predictors were calculated by averaging the response time, recognition accuracy, and reactivity ratings across all scenarios for each participant. Participants with incomplete submissions, submissions with repeated entries to open-ended questions, and submissions with responses unrelated to the question prompts were excluded from the final data analysis. The task was designed in a way that all responses were required. Therefore, no missing data was found. Three invalid responses from the rating task, which were indicated by untoggled Likert-scales, were treated as zeros in the final analysis.

#### Reliability

The Split-Half reliability was calculated to assess the internal consistency of the task at individual level. All 44 scenarios were split into two groups with 22 scenarios each. The number of scenarios from different consequence categories were matched between the halves and assigned into either group randomly. For each participant, all main variables were calculated for each half, and the correlations between the halves were then calculated using the Spearman–Brown formula to estimate reliability of the full task.

#### Main Model

A multi-step regression model with the percentage of risky choices as the outcome variable across individuals was constructed. Participants’ average degree of worry related to negative long-term consequences, approach-related motivation for positive short-term consequences, response time and accuracy for recognizing negative consequence prime words as a measure of implicit access to knowledge of long-term consequences were used as predictors.

Predictors were entered sequentially in the analysis. In the first step, indices of implicit access to long-term negative consequences (recognition accuracy and response time for recognizing prime words) were entered to assess their association with risky choices independent of explicit self-reported factors. In the second step, worry related to negative consequences was added to see if it accounted for additional variance in the model. In the last step, approach-related motivation for positive short-term outcomes was entered to assess its contribution in predicting percentage of risky choices.

#### Secondary and Exploratory Analysis

A logistic regression model with each participant’s “Yes” or “No” choice in each scenario as the binary outcome variable was constructed to explore associations with predictors on an individual-level. For exploratory analysis, a separate linear regression model with an individual’s BIS-11 score as the independent variable and percentage of risky choices as the dependent variable was made to probe the construct validity of the novel task as a measure of impulsivity.

## Results

### Sample Characteristics

After applying pre-registered exclusion criteria and excluding participants with repeated invalid responses, 40 participants were included in the final analysis. Because age was collected as a categorical variable, based on different categories’ midpoints, the mean age of the sample was estimated to be 38.01 years old. 72.5% of the participants felt more attracted to the image with more traditional female features. Full demographic information of the final 40 participants is shown in Table 1.

**Table 1.**
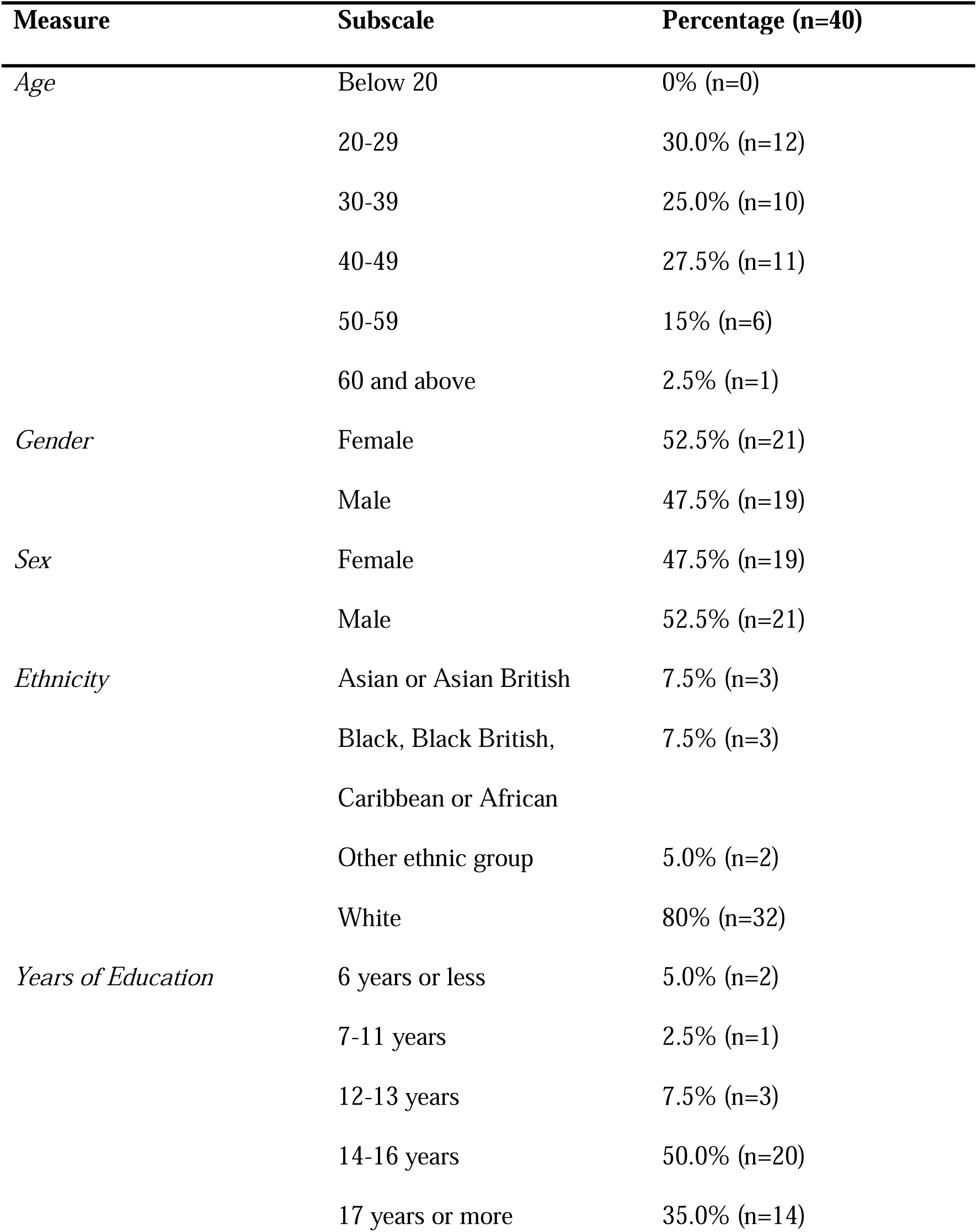
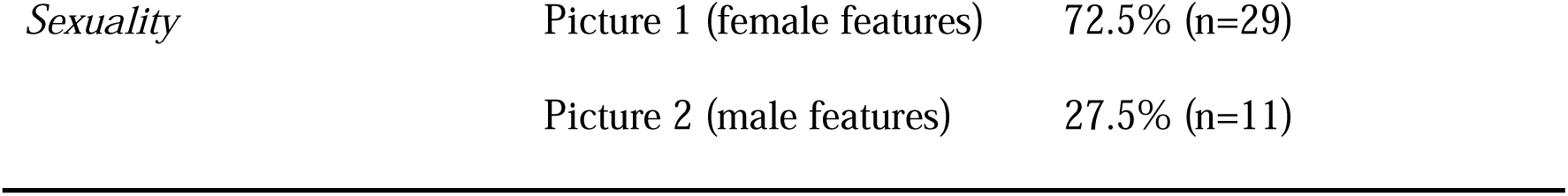
Demographic information of participants.

| **Measure** | **Subscale** | **Percentage (n=40)** |
| --- | --- | --- |
| *Age* | Below 20 | 0% (n=0) |
|  | 20-29 | 30.0% (n=12) |
|  | 30-39 | 25.0% (n=10) |
|  | 40-49 | 27.5% (n=11) |
|  | 50-59 | 15% (n=6) |
|  | 60 and above | 2.5% (n=1) |
| *Gender* | Female | 52.5% (n=21) |
|  | Male | 47.5% (n=19) |
| *Sex* | Female | 47.5% (n=19) |
|  | Male | 52.5% (n=21) |
| *Ethnicity* | Asian or Asian British | 7.5% (n=3) |
|  | Black, Black British,<br>Caribbean or African | 7.5% (n=3) |
|  | Other ethnic group | 5.0% (n=2) |
|  | White | 80% (n=32) |
| *Years of Education* | 6 years or less | 5.0% (n=2) |
|  | 7-11 years | 2.5% (n=1) |
|  | 12-13 years | 7.5% (n=3) |
|  | 14-16 years | 50.0% (n=20) |
|  | 17 years or more | 35.0% (n=14) |
| *Sexuality* | Picture 1 (female features) | 72.5% (n=29) |
|  | Picture 2 (male features) | 27.5% (n=11) |

### Variable Descriptives

Descriptive data for all primary variables are listed in Table 2. Participants made risky choices on an average of 43.35% of the trials (*SD* = 20.81%), indicating a substantial amount of variability in the number of impulsive choices made across individuals. The maximum percentage of impulsive choices was 100%, while the minimum was 13.64%. Participants’ ratings of approach-related motivation had a mean of 1.48 (*SD* = 0.71), whereas ratings of worry related to negative long-term consequences had a higher mean of 2.20 (*SD* = 0.76). Mean response time for recognizing negative consequence prime words was 10.74 seconds (*SD* = 4.90 s), while mean accuracy was 0.50 (*SD* = *0.54*).

**Table 2.**
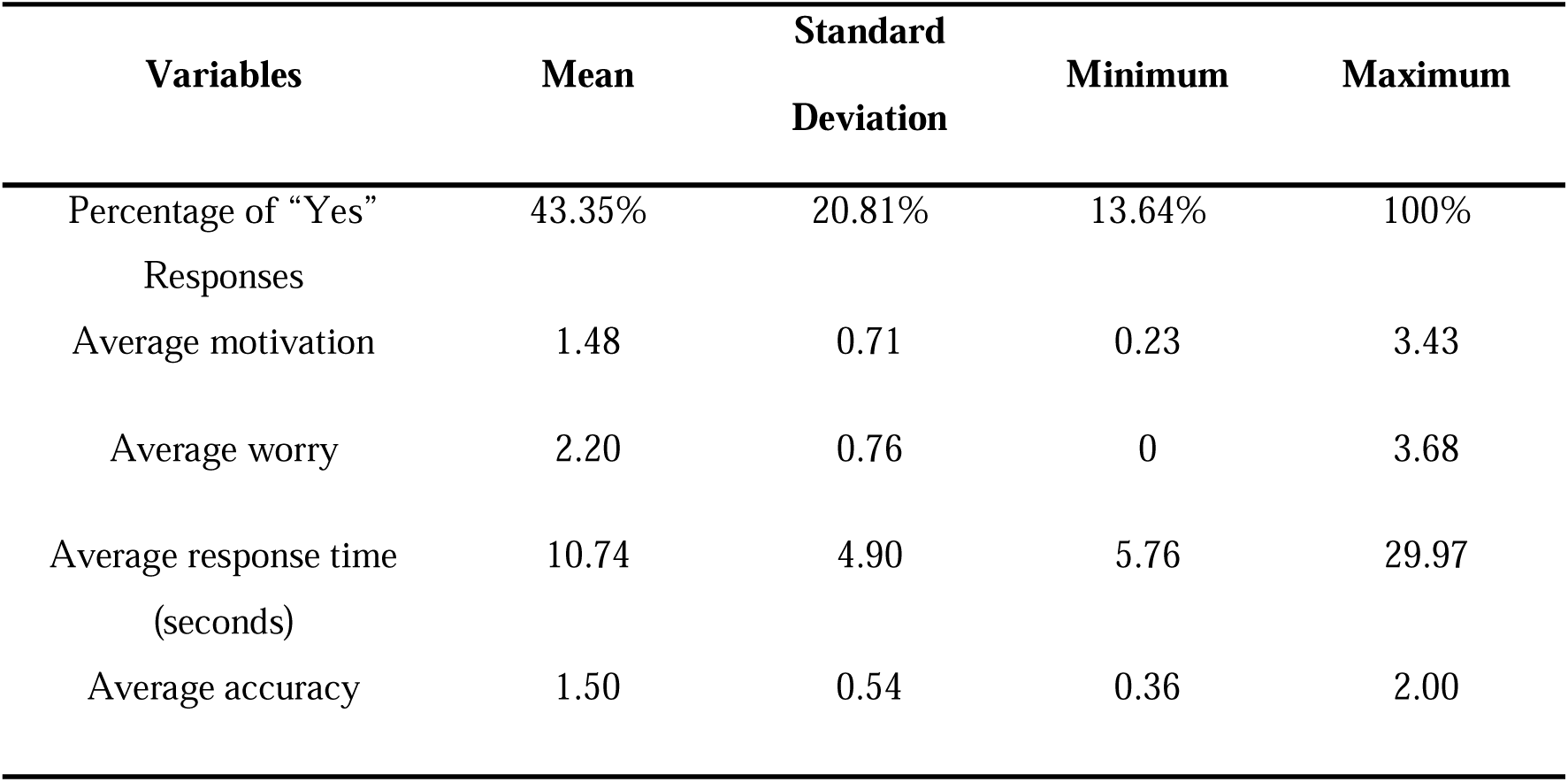
Descriptive statistics of all variables (n=40)

### Split-half Reliability

Split-half reliability of the task was estimated using the Spearman-Brown formula. The 44 scenarios were divided randomly into two matched halves of 22 scenarios each, while maintaining an approximately equal distribution of negative consequence categories across the halves. For each consequence category, scenarios were randomly assigned such that both halves contained the same number of items. For example, six scenarios involving addiction-related consequences were evenly distributed, with three scenarios allocated to each half. For each participant, main variables were computed separately for each half, and correlations were calculated across the halves. These correlations were adjusted using the Spearman–Brown formula. The resulting coefficients were 0.91 for the percentage of risky choices, 0.99 for consequence recognition accuracy, 0.93 for approach-related motivation, and 0.98 for worry related to negative consequences, indicating strong internal consistency in all primary variables across scenarios.

### Group-level Analysis

A multi-step regression analysis conducted in three steps was performed with the percentage of “yes” responses as the dependent variable as a measure of proneness to risky choices. The independent variables were each participant’s average worry related to negative long-term consequences, average approach-related motivation for positive short-term consequences, and average response time and accuracy in recognizing degraded auditory word stimuli. Results of the multiple regression analysis are listed in Table 3. In the first model, average accuracy and average response time were entered as predictors. The model explained 15.8% of the variance in percentage of “yes” responses (R^2^ =.158; Adjusted R² =.112). A significant negative relationship was found between average accuracy and percentage of “yes” responses, β = −.384, p = .018, such that higher accuracy in recognizing aurally degraded negative consequences stimuli was associated with a lower percentage of risky choices. In the second model, average worry was added. This model explained 18.2% of the variance (R² = .182; Adjusted R² = .114), reflecting only a small increase in explained variance (ΔR² = .024). Average accuracy remained a significant negative predictor, β = −.411, p = .013, whereas worry was not significant, β = .159, p = .304. Lastly, in the third model, average motivation related to positive short-term consequences was entered as a fourth predictor. The final model explained 57.8% of the variance in risky choices (R² = .578; Adjusted R² = .530), representing a substantial increase in explained variance (ΔR² = .396). In this model, approach-related motivation emerged as the strongest predictor, β = .700, t (35) = 5.73, p < .001, indicating that higher approach motivation was associated with a greater percentage of risky choices. After including motivation, accuracy, response time, and worry were no longer significant predictors.

**Table 3.** Multiple regression analysis predicting risky choices.

| Model | Unstandardized |  | Standardized | T | Sig. |
| --- | --- | --- | --- | --- | --- |
|  | Coefficients |  | Coefficients |  |  |
|  | B | Std. Error | Beta |  |  |
| Average accuracy | -0.148 | 0.060 | -0.384 | -2.477 | 0.018* |
| Average response time | 0.002 | 0.007 | 0.044 | 0.285 | 0.777 |
| Average accuracy | -0.158 | 0.060 | -0.411 | -2.617 | 0.013* |
| Average response time | 0.001 | 0.007 | 0.030 | 0.195 | 0.847 |
| Average worry | 0.044 | 0.042 | 0.159 | 1.042 | 0.304 |
| Average accuracy | -0.066 | 0.047 | -0.172 | -1.410 | 0.167 |
| Average response time | -4.828E<sup>-5</sup> | 0.005 | -0.001 | -0.010 | 0.992 |
| Average worry | -0.016 | 0.032 | -0.059 | -0.497 | 0.622 |
| Average motivation | 0.205 | 0.036 | 0.700 | 5.731 | <0.001*** |

Note. \*\*\*: $p < 0.001$ ; \*: $p < 0.05$ . $n = 40$ . Outcome variable was % of “yes” responses as a measure of proneness to risky choices.

### Additional Pre-registered Analysis

As a secondary analysis, a logistic regression model was constructed to predict trial-level “Yes” or “No” binary choices of impulsive decisions using worry and motivation ratings, response time, and response accuracy. However, this model could not be built reliably due to violations of model assumptions and lack of variation in both outcome and predictor variables in some participants. Three subjects chose 100% risky choices, resulting in no within-subject variance in the binary outcome, which in turn caused fitting problems in output. In addition, seven participants achieved full accuracy in prime word recognition, and one participant showed no worry towards any presented negative long-term consequences. As a result, those values were treated as constants and omitted by SPSS, and the model failed to converge. Therefore, this pre-registered model was not conducted in the end.

A linear regression model was constructed with percentage of “Yes” responses as the dependent variable and BIS total score as the independent variable. No significant relationship was detected between these two variables (B=0.185, *p*=0.245).

## Discussion

The present study developed a novel experimental task to disentangle cognitive components of risky choices, demonstrated excellent split-half reliability, and confirmed the hypothesis that weakened implicit access to knowledge of long-term negative consequences contributes to risky choices. To our knowledge, this is the first time this has been experimentally demonstrated. Participants who showed weaker access to long-term consequence-related information, as measured by recognizing the aurally degraded consequences, tended to endorse risky behaviors more frequently, which is in keeping with the prediction that decreased implicit access to representations of negative consequences is a possible component underlying impulsive behavior. This prediction was derived from research in patients with FTLD and frontopolar neurodegeneration who showed selective impairments of knowledge of long-vs. short-term consequences and social concepts (Zahn et al., 2017). Although more impulsivity-prone individuals without brain lesions are unlikely to lack this knowledge, they may nevertheless have greater difficulty accessing information about the consequences of social behavior or weigh negative outcomes differently when making impulsive decisions. Therefore, present findings provide preliminary support for weakened accessibility of long-term knowledge of negative consequences as an important cognitive process underlying impulsivity in a non-clinical sample.

Interestingly, response time as another implicit measure of knowledge accessibility showed no correlation with percentage of risky choices. One possible explanation for this absence considers the relative reliability of response time and accuracy-based measures in priming tasks. That is, accuracy-based measures have been shown in some contexts to exhibit higher reliability than reaction-time-base measures, and a shift from measuring reaction times to measuring accuracy to improve priming task reliability has been observed (Cameron et al., 2012; Draheim et al., 2021; Payne, 2005). Based on this perspective, the rejected hypothesis of a positive correlation between response time for recognizing prime words and percentage of risky choices does not necessarily undermine the relevance of consequence accessibility as a facet of impulsivity. Rather, it suggests that accuracy may be a more reliable indicator for implicit access of knowledge of long-term negative consequences in the RSC task than response time.

The hypothesis that higher approach-related motivation for positive short-term outcomes would be associated with a greater percentage of risky choices was supported. In fact, approach-related motivation emerged as the strongest predictor in the final multi-step regression model among all other predictors, accounting for substantial variance in the model. This finding is consistent with a large body of research conceptualizing impulsivity as heightened sensitivity to immediate rewards or incentives, especially in substance/alcohol use research (Dawe et al., 2004; Fromme et al., 1997; Martin & Potts, 2004). From a methodological point of view, this result demonstrates that the RSC task can successfully capture individual differences in approach-related motivation for immediate small rewards. Although the task does not involve explicit trade-offs between immediate and delayed rewards, heightened sensitivity to immediate reward is a core component in the delay discounting paradigm of impulsivity. For example, individuals with higher sensitivity for rewards showed deficits in choosing more rational options (Veldhoven et al., 2020).

The last variable in the multi-step regression model was worry related to negative long-term consequences, and no significantly negative association was found between it and risky choices. This could partially be explained by previous literature proposing that awareness of or explicit worry or concern about negative consequences does not necessarily lead to self-regulation or self-control, especially when immediate incentives are more salient, eliciting more intense emotions (Loewenstein et al., 2001; Metcalfe & Mischel, 1999).

Taken together, these findings suggest that impulsive decision-making may reflect the result of multiple cognitive processes, including motivational drive and accessibility of negative-consequence-related knowledge. This interpretation aligns with broader accounts of complex behavior as emerging from interactions among distributed cognitive systems (Barbey, 2018), and more specifically with models that conceptualize impulsivity as a multidimensional construct composed of distinct psychological and neural mechanisms (Dalley & Robbins, 2017).

### Methodological Contribution and Reliability

The current study developed and tested a novel experimental task intended to measure implicit access of knowledge of long-term negative consequences during impulsive decision-making. Split-half reliability estimates were substantially high for the primary behavioral outcome, as well as for motivation, worry, and prime word recognition accuracy and response time that served as measures of implicit access. This level of internal consistency is notable, given that a task with complex and heterogeneous design is often assumed to have lower reliability. Overall, the statistics support the use of the RSC task for reuse or validation in future experiments

### Secondary and Exploratory Analysis

A secondary analysis of a logistic regression model predicting trial-level binary outcome of impulsive choices was planned but could not be reported due to insufficient variability in both outcome and predictor variables for a small group of participants, resulting in a non-convergence issue for the model. Specifically, some participants achieved full accuracy on the prime word recognition task, while others consistently chose “Yes” responses or reported no worry towards any of the negative consequences. This result calls attention to practical implications for future research to consider. For example, individual-level modeling for larger samples and design modifications to avoid extreme response patterns. However, the constraints found in the secondary model should not weaken the primary finding and the reliability of the task.

An exploratory analysis examined the association between self-reported trait impulsivity, as measured by BIS-11, and the proportion of risky choices in the task. No significant relationship was observed between BIS total scores and risky choices. This finding is consistent with prior work demonstrating poor convergence between trait impulsivity and behavioral measures of impulsivity (Stahl et al., 2014). Instead of undermining the validity of the task, this dissociation provides further evidence that self-reports and behavioral tasks may capture distinct aspects of impulsivity, reiterating the importance of this study, which matches the aspect of interest with the correct tool that successfully assesses the intended aspect.

### Limitations and Future Directions

Several limitations are identified. Firstly, the limited sample size weakened statistical power for detecting smaller effects and interfered with the stability of multivariate models. Secondly, as mentioned before, current task design supported extreme response patterns, resulting in low variability in both outcome and predictor variables. Lastly, in the current study, only four factors were included in the model and examined. However, there are other factors that may also influence people’s impulsive choices, like the role of past experience. As shown in addiction research, the concept of “gateway hypothesis”, which states that an adolescent’s early experimentation with alcohol, tobacco, or cannabis (marijuana) may act as a gateway to more addictive and illicit substances when they grow older (Kandel & Kandel, 2014; Lynskey et al., 2003), suggests that past experience should also be taken into consideration when predicting impulsive choices. Therefore, for future research, more participants are needed to investigate the involvement of other factors in influencing people’s impulsive choices. Moreover, future research could apply this task to specific populations, such as patients with hypomania/mania or substance use disorder, to investigate the cognitive factors underpinning impulsive choices in psychopathology.

### Conclusions

In conclusion, the present study provides preliminary evidence that weakened accessibility of knowledge of long-term negative consequences can be seen as a potentially meaningful and measurable aspect of impulsivity. Results also provide strong support for the crucial role of approach-related motivation for short-term rewards in impulsive decision-making. Lastly, findings support using the RSC task as a reliable tool for measuring impulsive decision making and its component processes, but it could not be validated against a standard impulsivity scale and thus needs further validation against clinically relevant measures of impulse-related psychiatric disorders. Together, the study introduces a distinct aspect of impulsivity and a validated tool for investigating impulsivity as a multifaceted construct.

## Declaration

The authors would like to thank all participants for their time and engagement in the study. We are grateful to Dr. Andrew Lawrence for helpful discussions regarding statistical analyses.

HM is associated with the German Research Foundation (DFG)-funded International Research Training Group (IRTG) 2773 “Risk Factors and Pathomechanisms of Affective Disorders”. DF was funded by a King’s College London/IDOR Pioneer Science Fellowship. RZ was partly funded by MRC grant MR/Y008545/1 and National Institute for Health and Care Research (NIHR) Biomedical Research Centre at South London and Maudsley NHS Foundation Trust and King’s College London. The views expressed are those of the authors and not necessarily those of the NHS, the NIHR or the Department of Health and Social Care.

## Data Availability

All data produced are available online at https://osf.io/qt2mv/overview

## References

1. Barbey, A. K. (2018). Network neuroscience theory of human intelligence. Trends in Cognitive Sciences, 22(1), 8–20. 10.1016/j.tics.2017.10.001.

2. Bezdjian, S., Baker, L. A., Lozano, D. I., & Raine, A. (2009). Assessing inattention and impulsivity in children during the Go/NoGo task. The British Journal of Developmental Psychology, 27(Pt 2), 365–383. 10.1348/026151008X314919

3. Cameron, C. D., Brown-Iannuzzi, J. L., & Payne, B. K. (2012). Sequential priming measures of implicit social cognition: A meta-analysis of associations with behavior and explicit attitudes. *Personality and Social Psychology Review: An Official Journal of the Society for Personality and Social Psychology*, Inc, 16(4), 330–350. 10.1177/1088868312440047

4. Cyders, M. A., & Coskunpinar, A. (2011). Measurement of constructs using self-report and behavioral lab tasks: Is there overlap in nomothetic span and construct representation for impulsivity? Clinical Psychology Review, 31(6), 965–982. 10.1016/j.cpr.2011.06.001

5. Dalley, J. W., & Robbins, T. W. (2017). Fractionating impulsivity: Neuropsychiatric implications. Nature Reviews. Neuroscience, 18(3), 158–171. 10.1038/nrn.2017.8

6. Dawe, S., Gullo, M. J., & Loxton, N. J. (2004). Reward drive and rash impulsiveness as dimensions of impulsivity: Implications for substance misuse. Addictive Behaviors, 29(7), 1389–1405. 10.1016/j.addbeh.2004.06.004

7. Deyoung, C. (2010). Impulsivity as a personality trait. *Handbook of Self-Regulation: Research*, Theory, and Applications, 485–502.

8. Dimoska-Di Marco, A., McDonald, S., Kelly, M., Tate, R., & Johnstone, S. (2011). A meta-analysis of response inhibition and Stroop interference control deficits in adults with traumatic brain injury (TBI). Journal of Clinical and Experimental Neuropsychology, 33(4), 471–485. 10.1080/13803395.2010.533158

9. Draheim, C., Tsukahara, J. S., Martin, J. D., Mashburn, C. A., & Engle, R. W. (2021). A toolbox approach to improving the measurement of attention control. Journal of Experimental Psychology: General, 150(2), 242–275. 10.1037/xge0000783

10. ElevenLabs. (2024). *ElevenLabs* [Artificial intelligence voice generation software]. https://elevenlabs.io/

11. Eriksen, B. A., & Eriksen, C. W. (1974). Effects of noise letters upon the identification of a target letter in a nonsearch task. Perception & Psychophysics, 16(1), 143–149. 10.3758/BF03203267

12. Fromme, K., Katz, E. C., & Rivet, K. (1997). Outcome expectancies and risk-taking behavior. Cognitive Therapy and Research, 21(4), 421–442. 10.1023/A:1021932326716

13. Greenwald, A. G., Banaji, M. R., Rudman, L. A., Farnham, S. D., Nosek, B. A., & Mellott, D. S. (2002). A unified theory of implicit attitudes, stereotypes, self-esteem, and self-concept. Psychological Review, 109(1), 3–25. 10.1037/0033-295x.109.1.3

14. Grossman, M., Eslinger, P. J., Troiani, V., Anderson, C., Avants, B., Gee, J. C., McMillan, C., Massimo, L., Khan, A., & Antani, S. (2010). THE ROLE OF VENTRAL MEDIAL PREFRONTAL CORTEX IN SOCIAL DECISIONS: CONVERGING EVIDENCE FROM fMRI AND FRONTOTEMPORAL LOBAR DEGENERATION. Neuropsychologia, 48(12), 3505–3512. 10.1016/j.neuropsychologia.2010.07.036

15. Hamdan-Mansour, A. M., Mahmoud, K. F., Al Shibi, A. N., & Arabiat, D. H. (2018). Impulsivity and Sensation-Seeking Personality Traits as Predictors of Substance Use Among University Students. Journal of Psychosocial Nursing and Mental Health Services, 56(1), 57–63. 10.3928/02793695-20170905-04

16. Kandel, E. R., & Kandel, D. B. (2014). A Molecular Basis for Nicotine as a Gateway Drug. New England Journal of Medicine, 371(10), 932–943. 10.1056/NEJMsa1405092

17. Krueger, F., & Grafman, J. (2009). The medial prefrontal cortex mediates social event knowledge. Trends in Cognitive Sciences, 13(3), 103–109. 10.1016/j.tics.2008.12.005

18. Lee-Winn, A. E., Townsend, L., Reinblatt, S. P., & Mendelson, T. (2016). Associations of Neuroticism and Impulsivity with Binge Eating in a Nationally Representative Sample of Adolescents in the United States. Personality and Individual Differences, 90, 66–72. 10.1016/j.paid.2015.10.042

19. Leigh, B. C. (1999). Peril, chance, adventure: Concepts of risk, alcohol use and risky behavior in young adults. *Addiction (Abingdon*, England), 94(3), 371–383. 10.1046/j.1360-0443.1999.9433717.x

20. Li, Q., Zhao, Y., Gong, B., Li, R., Wang, Y., Yan, X., & Wu, C. (2022). Visual Affective Stimulus Database: A Validated Set of Short Videos. *Behavioral Sciences (Basel*, Switzerland), 12(5), 137. 10.3390/bs12050137

21. Loewenstein, G. F., Weber, E. U., Hsee, C. K., & Welch, N. (2001). Risk as feelings. Psychological Bulletin, 127(2), 267–286. 10.1037/0033-2909.127.2.267

22. Lynskey, M. T., Heath, A. C., Bucholz, K. K., Slutske, W. S., Madden, P. A. F., Nelson, E. C., Statham, D. J., & Martin, N. G. (2003). Escalation of drug use in early-onset cannabis users vs co-twin controls. JAMA, 289(4), 427–433. 10.1001/jama.289.4.427

23. MacPherson, L., Reynolds, E. K., Daughters, S. B., Wang, F., Cassidy, J., Mayes, L. C., & Lejuez, C. W. (2010). Positive and Negative Reinforcement Underlying Risk Behavior in Early Adolescents. Prevention SciencelJ: The Official Journal of the Society for Prevention Research, 11(3), 331–342. 10.1007/s11121-010-0172-7

24. Martin, L. E., & Potts, G. F. (2004). Reward sensitivity in impulsivity. Neuroreport, 15(9), 1519–1522. 10.1097/01.wnr.0000132920.12990.b9

25. Metcalfe, J., & Mischel, W. (1999). A hot/cool-system analysis of delay of gratification: Dynamics of willpower. Psychological Review, 106(1), 3–19. 10.1037/0033-295x.106.1.3

26. Moll, J., Zahn, R., de Oliveira-Souza, R., Krueger, F., & Grafman, J. (2005). Opinion: The neural basis of human moral cognition. Nature Reviews. Neuroscience, 6(10), 799–809. 10.1038/nrn1768

27. O’Reilly, J. M., Hubbard, M. L., Lessler, J. T., Biemer, P. P., & Turner, C. F. (1994). AUDIO AND VIDEO COMPUTER-ASSISTED SELF INTERVIEWING. Journal of Official Statistics, 10(2), 197–214.

28. Patton, J. H., Stanford, M. S., & Barratt, E. S. (1995). Factor structure of the Barratt impulsiveness scale. Journal of Clinical Psychology, 51(6), 768–774. 10.1002/1097-4679(199511)51:6<768::aid-jclp2270510607>3.0.co;2-1

29. Payne, B. K. (2005). Conceptualizing control in social cognition: How executive functioning modulates the expression of automatic stereotyping. Journal of Personality and Social Psychology, 89(4), 488–503. 10.1037/0022-3514.89.4.488

30. Reise, S. P., Moore, T. M., Sabb, F. W., Brown, A. K., & London, E. D. (2013). The Barratt Impulsiveness Scale - 11: Reassessment of its Structure in a Community Sample. Psychological Assessment, 25(2), 631–642. 10.1037/a0032161

31. Roberts, B. W., & Mroczek, D. (2008). Personality Trait Change in Adulthood. Current Directions in Psychological Science, 17(1), 31–35. 10.1111/j.1467-8721.2008.00543.x

32. Runway. (2024). *Gen-2* [Artificial intelligence video generation software]. https://runwayml.com/

33. Scarborough, D. L., Cortese, C., & Scarborough, H. S. (1977). Frequency and repetition effects in lexical memory. Journal of Experimental Psychology: Human Perception and Performance, 3(1), 1–17. 10.1037/0096-1523.3.1.1

34. Schacter, D. L., & Buckner, R. L. (1998). Priming and the Brain. Neuron, 20(2), 185–195. 10.1016/S0896-6273(00)80448-1

35. Schacter, D. L., Dobbins, I. G., & Schnyer, D. M. (2004). Specificity of priming: A cognitive neuroscience perspective. Nature Reviews. Neuroscience, 5(11), 853–862. 10.1038/nrn1534

36. Stahl, C., Voss, A., Schmitz, F., Nuszbaum, M., Tüscher, O., Lieb, K., & Klauer, K. C. (2014). Behavioral components of impulsivity. Journal of Experimental Psychology. General, 143(2), 850–886. 10.1037/a0033981

37. Swann, A. C., Dougherty, D. M., Pazzaglia, P. J., Pham, M., Steinberg, J. L., & Moeller, F. G. (2005). Increased impulsivity associated with severity of suicide attempt history in patients with bipolar disorder. The American Journal of Psychiatry, 162(9), 1680–1687. 10.1176/appi.ajp.162.9.1680

38. Vasconcelos, A. G., Malloy-Diniz, L., & Correa, H. (2012). Systematic review of psychometric proprieties of Barratt Impulsiveness Scale Version 11 (BIS-11). Clinical Neuropsychiatry: Journal of Treatment Evaluation, 9(2), 61–74.

39. Veldhoven, D. T., Roozen, H., & Vingerhoets, A. (2020). The Association between Reward Sensitivity and Activity Engagement: The Influence of Delay Discounting and Anhedonia. Alcohol and Alcoholism, 55(2), 215–224. 10.1093/alcalc/agz105

40. Zahn, R., de Oliveira-Souza, R., & Moll, J. (2020). Moral Motivation and the Basal Forebrain. Neuroscience & Biobehavioral Reviews, 108, 207–217. 10.1016/j.neubiorev.2019.10.022

41. Zahn, R., Green, S., Beaumont, H., Burns, A., Moll, J., Caine, D., Gerhard, A., Hoffman, P., Shaw, B., Grafman, J., & Lambon Ralph, M. A. (2017). Frontotemporal lobar degeneration and social behaviour: Dissociation between the knowledge of its consequences and its conceptual meaning. Cortex; a Journal Devoted to the Study of the Nervous System and Behavior, 93, 107–118. 10.1016/j.cortex.2017.05.009

